# Preliminary report: Safety and immunogenicity of an inactivated SARS-CoV-2 vaccine, KD-414, in healthy adult participants: a non-randomized, open-label phase 2/3 clinical study in Japan

**DOI:** 10.1101/2022.10.27.22281603

**Authors:** Keishi Kido, Kayo Ibaragi, Mitsuyoshi Tanishima, Yosuke Muramoto, Shun Nakayama, Kohei Ata, Kenshi Hayashida, Hideki Nakamura, Yasuhiko Shinmura, Yoshiaki Oda, Masafumi Endo, Kengo Sonoda, Yuji Sasagawa, Yasuhiro Iwama, Kohji Ueda, Takayuki Matsumoto

## Abstract

**Background:** In the prolonged COVID-19 pandemic, there remains a high need for the development of a severe acute respiratory syndrome coronavirus 2 (SARS-CoV-2) vaccine that can be used more safely and effectively to prevents the disease onset or severe disease. To satisfy such unmet need, we are currently developing the inactivated whole particle SARS-CoV-2 vaccine (KD-414) and conducted a phase 2/3 study in healthy adults in Japan to accumulate more immunogenicity and safety data of KD-414 using the dose selected based on the results of the phase 1/2 study.

**Methods:** In an open-label uncontrolled phase 2/3 study, adults aged 18 years or older without a history of COVID-19 or COVID-19 vaccination received two intramuscular doses of KD-414 at a 28-day intervals, followed by one intramuscular dose 13 weeks after the second dose as the primary immunization. Safety data were collected after the first dose of KD-414 in all participants to evaluate the safety profile. In predetermined immunogenicity analysis subjects, the neutralizing antibody titers against the pseudovirus SARS-CoV-2 (Wuhan) before the first vaccination and after each vaccination with KD-414 were evaluated.

**Results:** A total of 2500 adults aged 18 years or older were enrolled; 2474 of them received the vaccination up to the second dose, and 2081 completed the third vaccination. Regarding the safety, no deaths or serious adverse reactions were recorded from the first vaccination until 28 days after the third vaccination with KD-414. The incidence of adverse reactions (number of participants with onsets/number of participants in the safety analysis set) was 80.6% (2015/2500). Adverse reactions with an incidence of 10% or more included injection site pain, malaise, headache, injection site erythema, myalgia, and injection site induration. A total of 11 events of grade 3 or higher adverse reactions that prevented daily activities in 9 participants. There was no increasing tendency in the incidence of adverse reactions responding to the vaccinations. To evaluate immunogenicity, 295 first comers enrolled from five age ranges were allocated to the immunogenicity analysis subjects; 291 participants received the vaccination up to the second dose, and 249 participants completed the third vaccination. The geometric mean titers (95% confidence interval [CI]) of neutralizing antibody titers against pseudovirus SARS-CoV-2 (Wuhan) 28 days after the second vaccination and 28 days after the third vaccination with KD-414 were 139.6 (118.9 - 164.0) and 285.6 (244.3 – 334.0), respectively, showing an approximately two-fold increase after the third vaccination compared to that after the second vaccination. The geometric mean titers (95% CI) of neutralizing antibody titers after the third vaccination were 327.6 (269.8 – 397.9), 272.2 (211.5 - 350.4) and 128.0 (51.6 - 317.7) in participants aged 18 to 40 years, 41 to 64 years, and 65 years or older, respectively, showing an age-dependency.

**Conclusion:** This study confirmed the favorable safety profile of KD-414 as a result of three vaccinations of KD-414 administered to over 2000 healthy Japanese participants aged 18 years or older. There were no particular differences in the types and incidences of adverse reactions between vaccinations, and no tendency of an increase in adverse reactions with an increase in the number of vaccinations. Similar to the phase 1/2 study, neutralizing antibody responses appeared to be age-dependent and the highest titers were observed in the age group of 18 - 40 years. A phase 3 study in adults aged 18 - 40 years (jRCT2031210679) and a phase 2/3 study in children aged 6 months - 18 years (jRCT2031220032) are currently ongoing.

## Introduction

COVID-19 is an infectious disease caused by severe acute respiratory syndrome coronavirus 2 (SARS-CoV-2), which was identified in December 2019 in Wuhan, Hubei Province, China. Thereafter, a Public Health Emergency of International Concern (PHEIC) was declared by the World Health Organization (WHO) on January 30, 2020 and a global pandemic was declared on March 11, 2020 ^1^. As of September 9, 2022, 610 million cases of SARS-CoV-2 infection have been reported, resulting in 6.5 million deaths, so vaccine development against COVID-19 remains a significant global challenge ^2^.

At present, vaccines against COVID-19, including inactivated vaccines, DNA vaccines and mRNA vaccines, viral vector vaccines, recombinant protein vaccines, etc. have been developed in the United States, the United Kingdom, China, and other countries around the world. In Japan, five products of mRNA vaccines, a viral vector vaccine, and a recombinant protein vaccine have been approved as of September 2022 ^3-7^. However, most of them are imported and there are concerns regarding adverse reactions associated with new modality, such as myocarditis and thrombosis with thrombocytopenia syndrome ^8-11^. Because of that, booster shots (a third dose and fourth dose) and primary vaccination in children who require higher safety are not progressing. It is important to secure sufficient amounts of vaccines against SARS-CoV-2 in Japan for future emergencies and, similar to seasonal influenza vaccines, these vaccines will be continuously required in the future. Thus, domestic vaccines with an effectiveness and high safety are needed.

KD-414 is a purified, inactivated, whole-virus SARS-CoV-2 vaccine, which is a modality that has been effective as multiple infectious disease vaccines and has an established safety. The manufacturing method is described in a previous report ^12^.

In the phase 1/2 study of KD-414, safety and immunogenicity were evaluated after two intramuscular vaccinations at three different doses of KD-414 which have different active ingredients (converted to protein amount): product L (2.5 μg/dose), product M (5 μg/dose), and product H (10 μg/dose). The results showed that the investigational product was well tolerated at all the dose levels, and the neutralizing antibody titer was the highest in the product H (10 μg/dose) group ^12^. Therefore, we decided to use product H (10 μg/dose) in the late-stage clinical studies of KD-414. However, although the preventive effect of the novel corona vaccine against the onset has been shown to correlate highly with the neutralizing antibody levels ^13^, the threshold of neutralizing antibody titers required for prevention has not yet been established. Therefore, it is important to evaluate the vaccination regimen of the three doses as a primary series that can induce higher neutralizing antibody titers.

Here, we report safety and immunogenicity findings from the phase 2/3 study conducted to evaluate the safety and immunogenicity of three doses of KD-414 in adults aged 18 years or older to accumulate more safety and immunogenicity data of KD-414 using the dose selected in the phase 1/2 study.

## Materials and Methods

### Study design, and participants

This multicenter, open-label, and uncontrolled study was conducted at 12 sites in Japan. The target number of participants was 2000, with healthy Japanese participants aged 18 years or older.

The study excluded participants with a history of COVID-19 or vaccination against COVID-19, including unapproved vaccines. In addition, participants confirmed to be infected with SARS-CoV-2 during the study were withdrawn from the study. During the trial, the investigator instructed the participants to undergo laboratory tests, including PCR, if the participant experienced fever, chills, cough, shortness of breath, and/or dyspnea for at least 24 hours or if the participant experienced malaise, myalgia, arthralgia, headache, dysgeusia, smell disorder, sore throat, stuffy nose, runny nose, nausea, vomiting, and diarrhea for at least 48 hours.

This study was reviewed and approved by the institutional review boards (IRBs) of all the institutions. Written informed consent was obtained from all participants before the screening. In addition, the study was conducted in compliance with the ethical principles of the Declaration of Helsinki, Good Clinical Practice (GCP) ordinance, etc., and registered in a jRCT at the initiation of the study (jRCT2071210081).

### Trial procedure

This study used KD-414 with 10 μg/dose of the active ingredients (converted to total protein amount) which was manufactured following good manufacturing practice (GMP) standards for investigational new drugs in Japan. A total of three vaccinations were administered for the primary series: two intramuscular vaccinations of KD-414 at an interval of 28 days followed by one intramuscular vaccination administered 13 weeks after the second vaccination. The original plan was to administer two doses at the start of this study. However, the plan was revised, and three doses were administered for the primary series during this study based on the results of the preceding phase 1/2 study. The change in the protocol was explained to all participants continuing on the study to obtain informed consent. The third vaccination was administered to participants who wished to receive the third vaccination among those who gave written informed consent.

Each participant recorded his/her health status and body temperature in an e-diary during the period beginning after each vaccination with KD-414 to day six after vaccination (a total of seven days, including the day of vaccination). The participants underwent a follow-up examination 28 days after the third vaccination. After completion of the follow-up examination, the participants entered a one-year follow-up period. participants who did not receive the second or third vaccination also entered a one-year follow-up period after the last vaccination.

For immunogenicity evaluation, up to 100, 50, 50, 50, and 50 participants were allocated to the immunogenicity analysis subjects by age groups of 18 - 29 years, 30 - 39 years, 40 - 49 years, 50 - 59 years, and 60 years or older, respectively, in the order of enrollment at pre-specified medical institutions. In immunogenicity analysis subjects, blood was collected before and 28 days after each dose and 13, 26, and 52 weeks after the third dose to measure the neutralizing antibody titer against the pseudovirus SARS-CoV-2 (Wuhan).

Injection site erythema, swelling, induration, and pain, which were observed after administration of each dose of KD-414 up to six days after vaccination, were defined as solicited local adverse events and were generally considered to be related to the investigational product. Fever, headache, malaise, nausea, and myalgia, which were observed after administration of each dose of KD-414 up to six days after vaccination, were defined as solicited systemic adverse events. All events not that could not be characterized as solicited adverse events were recorded as unsolicited adverse events. Adverse events were classified into five grades (grades 0 to 4). The criteria were in accordance with those of the Food and Drug Administration (FDA) ^14^, however, erythema/redness, induration/swelling, and fever with a severity of less than grade 1 were characterized as grade 0. Among all adverse events, those for which there was at least a reasonable possibility of a relationship with the investigational product that could not be ruled out were defined as adverse reactions.

The primary endpoint was the geometric mean neutralizing antibody titers (GMTs) of KD-414 against the pseudovirus SARS-CoV-2 (Wuhan) 28 days after the second vaccination and third vaccinations, respectively. As a secondary endpoint, the neutralizing antibody-seroconversion rates 28 days after the second and third vaccination with the investigational product, respectively, were evaluated. The neutralizing antibody-seroconversion rate was defined as the rate of participants whose neutralizing antibody titer increased at least 4-fold compared to that before the first vaccination of the investigational product.

### Pseudovirus SARS-CoV-2 (Wuhan) spike protein neutralizing antibody titers

The pseudovirus neutralizing antibody assay^12^ was performed at LabCorp Drug Development. The pseudovirus neutralizing antibody assay with a lentivirus-based pseudovirus particle expressing the SARS-CoV-2 (Wuhan) spike protein was validated at Monogram Biosciences and then transferred to LabCorp Drug Development. Briefly the mixed pseudoviruses and serially diluted serum samples were incubated in HEK293 cells expressing the ACE2 receptor. After incubation, the luciferase produced was measured to determine the pseudovirus neutralizing antibody titers (50% inhibition dose [ID50]) using the following calculation formula. The lower limit of quantitation (LLOQ) for pseudovirus neutralizing antibodies was 40 (ID50).

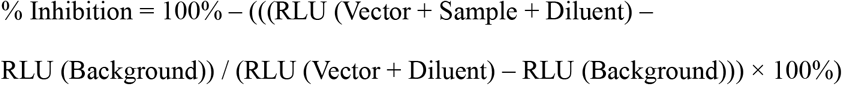

### Statistical Analysis

The target sample size for this study was set to 2000 participants to evaluate the safety profile of KD-414. Since this was an exploratory study, no hypothesis testing was performed with statistically powered sample sizes. Therefore, no adjustment for the multiplicity of type I errors was performed. In all the analyses, missing data were not imputed.

Safety analyses were performed for the KD-414-vaccinated population with evaluable safety data. The number of participants and incidence of solicited local adverse reactions, solicited systemic adverse reactions, and unsolicited adverse reactions were calculated.

Immunogenicity analysis was performed in the full analysis set (FAS) and per-protocol set (PPS). The results of the PPS analysis were handled as the primary results. The FAS involved the population of participants vaccinated with KD-414 among the immunogenicity analysis subjects, excluding those without data on neutralizing antibody titers after investigational product vaccination and those with deviation from GCP. PPS was defined as the population in which participants with major deviations from the protocol (related to the inclusion criteria, dosage and administration, prohibited concomitant medications/therapies, and timing of blood collection) were excluded from the FAS. For the primary evaluation, the GMTs were analyzed 28 days after the second and third vaccination, respectively, and the 95% CI was calculated based on the t-distribution. For the secondary evaluation, the seroconversion rate of the neutralizing antibody titer and the Clopper-Pearson 95% CI were calculated. In addition, subgroup analyses were performed according to the age group.

For other demographic characteristics factors, descriptive statistics were calculated for continuous data, and the frequencies and the percentages were calculated for categorical data.

SAS Release 9.4 (SAS Institute Inc., Cary, NC, USA) was used for the statistical analysis.

## Results

### Population

While the target number of participants was 2000, 2500 participants received the first vaccination of the investigational product, and 2474 participants received up to the second vaccination. A total of 150 participants discontinued the study after the second vaccination. Of these, 106 participants discontinued the study after they were confirmed to have been infected with SARS-CoV-2, according to the protocol. Written consent was obtained from 2324 participants for changes in protocol for the administration of additional third vaccination. The third vaccination was administered to 2081 participants who wished to receive the third vaccination. A total of 243 participants who did not wish to receive the third vaccination proceeded to the follow-up period after the second vaccination. Similarly, of the 295 participants in the immunogenicity analysis subjects, 291 participants received up to the second vaccination, and 249 participants received the third vaccination. None of the 2500 participants who received at least one dose of the investigational product were excluded from the safety set. Of the 295 participants in the immunogenicity analysis subjects, 292 participants were included in the FAS, of whom 287 participants were included in the PPS. (Figure 1)

**Figure 1:**
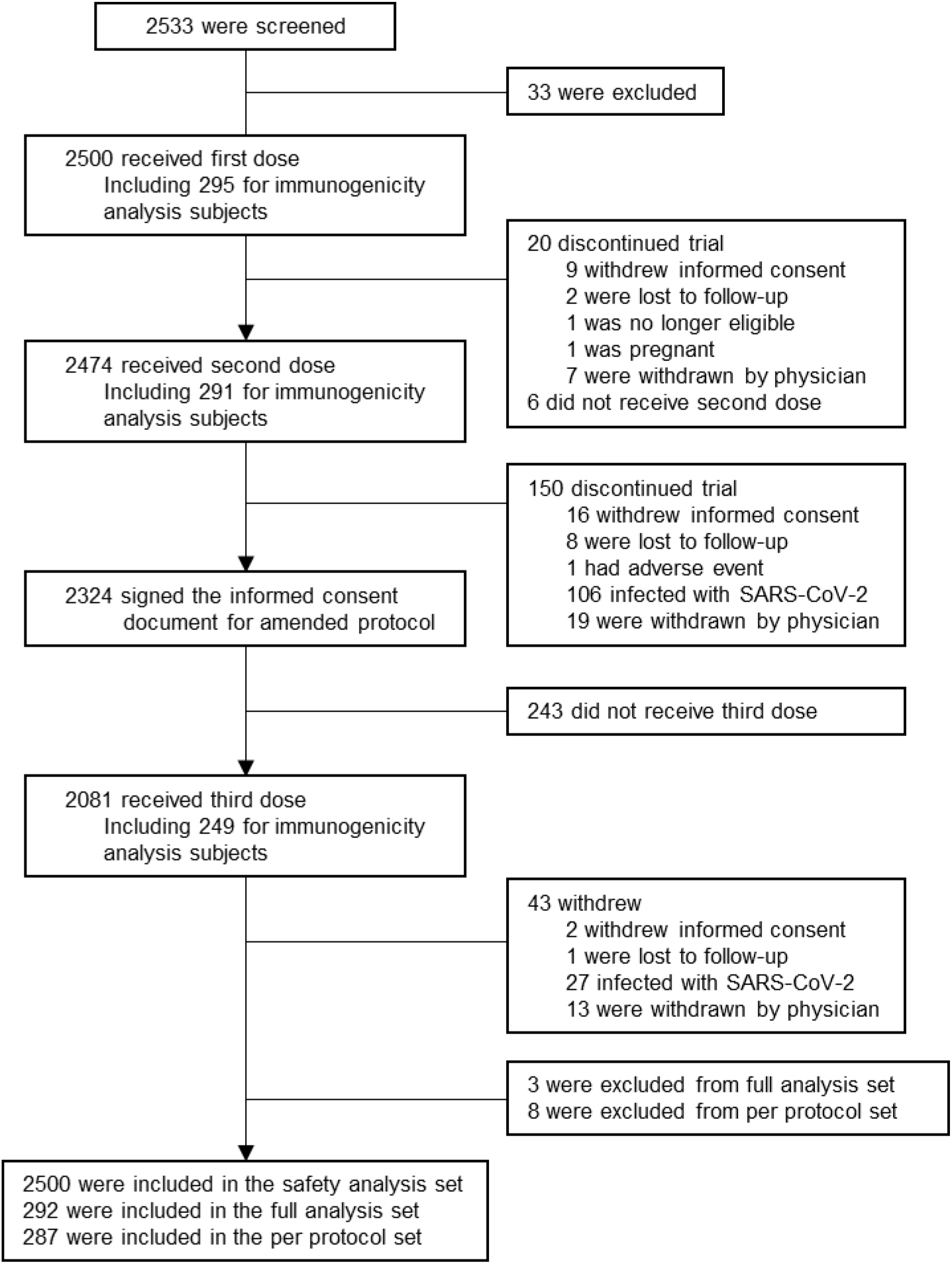
Trial profile.

The mean age of the participants in the safety analysis set was 43.2 years, with no particular preference for sex. The percentage of participants with the underlying diseases was 5.5%. The distributions of age, sex, and underlying diseases in the PPS were also similar. The percentage of participants with a negative neutralizing antibody titer against the pseudovirus SARS-CoV-2 before the first vaccination with the investigational product was 96.5%. (Table 1)

**Table 1:** Characteristics of the Participants

| Characteristic | Safety analysis set (N=2500) | Per protocol set (N=287) |
| --- | --- | --- |
| Age, years | 43.2(11.9) | 41.0(14.7) |
| 18-40 years | 1005(40.2) | 148(51.6) |
| 40-64 years | 1427(57.1) | 125(43.6) |
| ≥65 years | 68(2.7) | 14(4.9) |
| Sex |  |  |
| Male | 1200(48.0) | 144(50.2) |
| Female | 1300(52.0) | 143(49.8) |
| Underlying disease |  |  |
| With | 2363(94.5) | 269(93.7) |
| Without | 137(5.5) | 18(6.3) |
| Baseline anti-SARS CoV-2 neutralizing antibody |  |  |
| Negative | NA | 277(96.5) |
| Positive | NA | 10(3.5) |

Data are mean(SD) or n(%).
Underlying disease: such as cardiovascular diseases, renal diseases, hepatic diseases, hematological diseases, developmental disorders, respiratory diseases, or diabetes mellitus.

### Safety

No participants died. Four serious adverse events (SAE) other than death were observed in four participants during the period from after the first vaccination with the investigational product to 28 days after the second vaccination with the investigational product and the period from after the third vaccination to 28 days after the third vaccination. The investigator did not consider the SAE to be related to the investigational product. Nine events in seven participants required or should have led to discontinuation of the investigational product (significant adverse events). The investigator considered all significant adverse events related to the investigational product.

The incidence (number of participants with onset/number of participants in the safety analysis set) of adverse reactions from after the first vaccination to 28 days after the third vaccination of the investigational product in the safety analysis set, in which KD-414 was administered at least once, was 80.6% (2015/2500). The most common adverse reaction was injection site pain. Relatively common adverse reactions with an incidence of 10% or higher included injection site pain, malaise, headache, injection site erythema, myalgia, and injection site induration. When the incidence of adverse reactions was examined based on age, the incidence of solicited systemic adverse reactions was higher in participants aged 18 - 40 years than in participants aged 41 - 64 years or those aged 65 years or older. No noteworthy trends was observed in the incidence of adverse reactions in participants with or without underlying diseases. (Table 2)

**Table 2:**
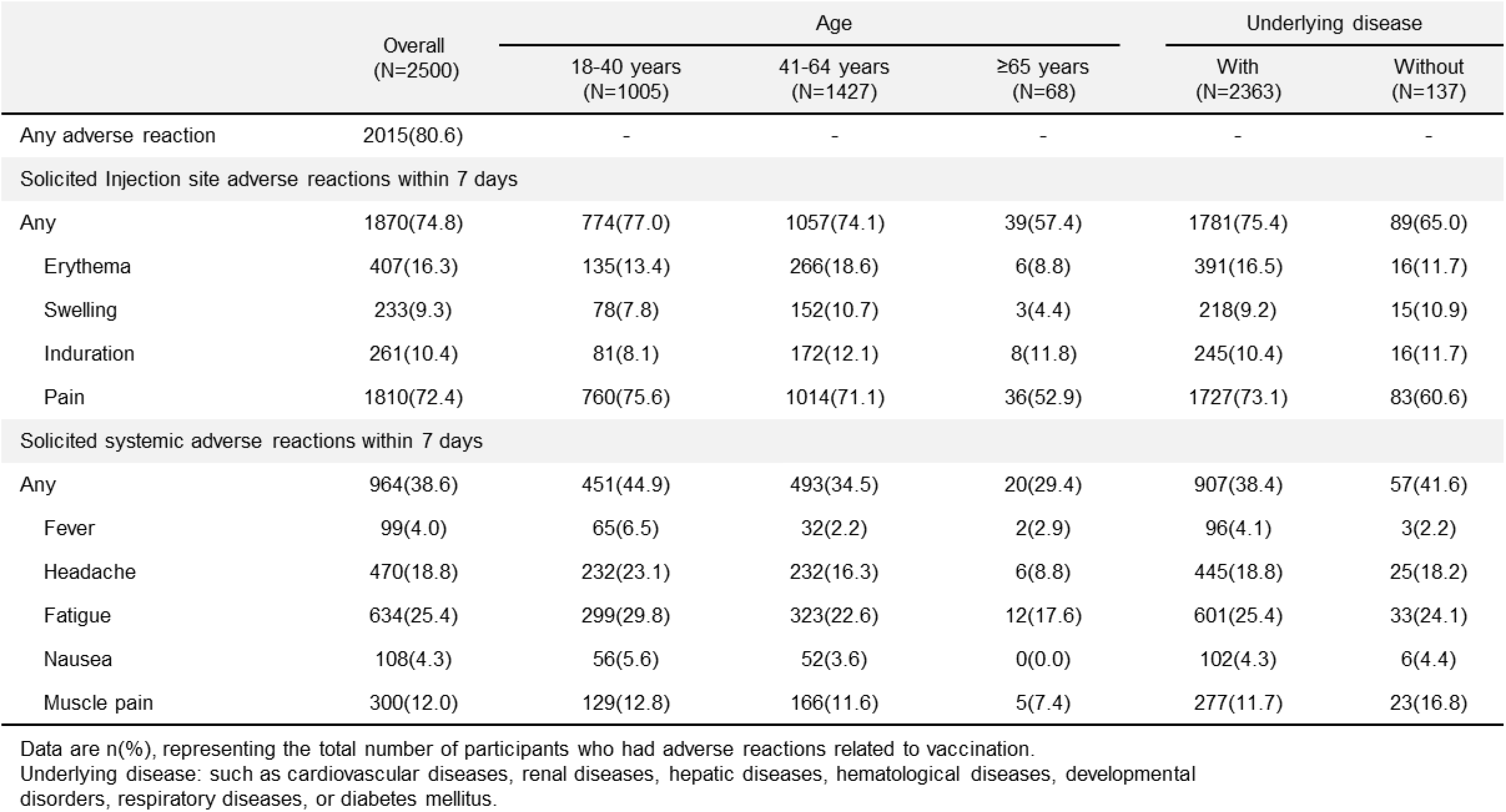
Adverse reactions within 7 days after the first, second, or third doses.

|  | Overall<br>(N=2500) | Age |  |  | Underlying disease |  |
| --- | --- | --- | --- | --- | --- | --- |
|  |  | 18-40 years<br>(N=1005) | 41-64 years<br>(N=1427) | ≥65 years<br>(N=68) | With<br>(N=2363) | Without<br>(N=137) |
| Any adverse reaction | 2015(80.6) | - | - | - | - | - |
| Solicited Injection site adverse reactions within 7 days |  |  |  |  |  |  |
| Any | 1870(74.8) | 774(77.0) | 1057(74.1) | 39(57.4) | 1781(75.4) | 89(65.0) |
| Erythema | 407(16.3) | 135(13.4) | 266(18.6) | 6(8.8) | 391(16.5) | 16(11.7) |
| Swelling | 233(9.3) | 78(7.8) | 152(10.7) | 3(4.4) | 218(9.2) | 15(10.9) |
| Induration | 261(10.4) | 81(8.1) | 172(12.1) | 8(11.8) | 245(10.4) | 16(11.7) |
| Pain | 1810(72.4) | 760(75.6) | 1014(71.1) | 36(52.9) | 1727(73.1) | 83(60.6) |
| Solicited systemic adverse reactions within 7 days |  |  |  |  |  |  |
| Any | 964(38.6) | 451(44.9) | 493(34.5) | 20(29.4) | 907(38.4) | 57(41.6) |
| Fever | 99(4.0) | 65(6.5) | 32(2.2) | 2(2.9) | 96(4.1) | 3(2.2) |
| Headache | 470(18.8) | 232(23.1) | 232(16.3) | 6(8.8) | 445(18.8) | 25(18.2) |
| Fatigue | 634(25.4) | 299(29.8) | 323(22.6) | 12(17.6) | 601(25.4) | 33(24.1) |
| Nausea | 108(4.3) | 56(5.6) | 52(3.6) | 0(0.0) | 102(4.3) | 6(4.4) |
| Muscle pain | 300(12.0) | 129(12.8) | 166(11.6) | 5(7.4) | 277(11.7) | 23(16.8) |

Data are n(%), representing the total number of participants who had adverse reactions related to vaccination. Underlying disease: such as cardiovascular diseases, renal diseases, hepatic diseases, hematological diseases, developmental disorders, respiratory diseases, or diabetes mellitus.

Eleven adverse reactions of high severity (grade 3 or higher) were observed in nine participants. Most adverse reactions were of grade 1 or 2. (Table 3)

**Table 3:** Severity of Adverse reactions within 7 days after the first, second, or third doses.

| Overall(N=2500) | Grade 0 | Grade 1 | Grade 2 | Grade 3 |
| --- | --- | --- | --- | --- |
| Solicited Injection site adverse reactions within 7 days |  |  |  |  |
| Any | 54(2.2) | 1729(69.2) | 86(3.4) | 1(0.0) |
| Erythema | 392(15.7) | 12(0.5) | 3(0.1) | 0(0.0) |
| Swelling | 182(7.3) | 44(1.8) | 7(0.3) | 0(0.0) |
| Induration | 216(8.6) | 37(1.5) | 8(0.3) | 0(0.0) |
| Pain | NA | 1735(69.4) | 74(3.0) | 1(0.0) |
| Solicited systemic adverse reactions within 7 days |  |  |  |  |
| Any | 22(0.9) | 827(33.1) | 108(4.3) | 7(0.3) |
| Fever | 79(3.2) | 13(0.5) | 5(0.2) | 2(0.1) |
| Headache | NA | 421(16.8) | 47(1.9) | 2(0.1) |
| Fatigue | NA | 563(22.5) | 68(2.7) | 3(0.1) |
| Nausea | NA | 98(3.9) | 9(0.4) | 1(0.0) |
| Muscle pain | NA | 282(11.3) | 17(0.7) | 1(0.0) |

Data are n/N(%), representing the total number of participants who had adverse reactions related to vaccination.
The severity of adverse events were graded as Grade0, Grade1 (Mild), Grade2 (Moderate), Grade3 (Severe), and Grade4 (Potentially life threatening).

No differences were observed in the types of adverse reactions associated with first, second and third vaccinations, and no noteworthy tendency was observed with respect to the incidences. (Table 4) From the first vaccination with the investigational product to the completion of the follow-up examination, no case of suspected vaccine-associated enhanced respiratory disease (ERD) or antibody dependent enhancement (ADE) was reported.

**Table 4:** Solicited and unsolicited adverse reactions within 7 days after each dose.

|  | After 1<sup>st</sup> dose<br>(N=2500) | After 2<sup>nd</sup> dose<br>(N=2474) | After 3<sup>rd</sup> dose<br>(N=2081) |
| --- | --- | --- | --- |
| Solicited Injection site adverse reactions within 7 days |  |  |  |
| Any | 1344(53.8) | 1407(56.9) | 1109(53.3) |
| Erythema | 178(7.1) | 214(8.6) | 201(9.7) |
| Swelling | 65(2.6) | 112(4.5) | 115(5.5) |
| Induration | 84(3.4) | 125(5.1) | 123(5.9) |
| Pain | 1266(50.6) | 1346(54.4) | 1039(49.9) |
| Solicited systemic adverse reactions within 7 days |  |  |  |
| Any | 563(22.5) | 526(21.3) | 393(18.9) |
| Fever | 43(1.7) | 38(1.5) | 33(1.6) |
| Headache | 243(9.7) | 243(9.8) | 159(7.6) |
| Fatigue | 351(14.0) | 319(12.9) | 269(12.9) |
| Nausea | 48(1.9) | 50(2.0) | 30(1.4) |
| Muscle pain | 160(6.4) | 123(5.0) | 90(4.3) |
| Unsolicited systemic adverse reactions within 28 days |  |  |  |
| Any | 294(11.8) | 225(9.1) | 147(7.1) |

Data are n(%), representing the total number of participants who had adverse reactions related to vaccination.

### Neutralizing responses

The GMTs (95% CI) for pseudovirus SARS-CoV-2 28 days after the second and third vaccination of the investigational product were 139.6 (118.9 - 164.0) and 285.6 (244.3 – 334.0), respectively, showing an approximately two-fold increase after the third vaccination compared to that after the second vaccination. The neutralizing antibody-seroconversion rates (95% CI) 28 days after the second and third vaccination were 67.0% (61.2% - 72.5%) and 84.5% (79.3% - 88.9%), respectively. (Table 5)

**Table 5:**
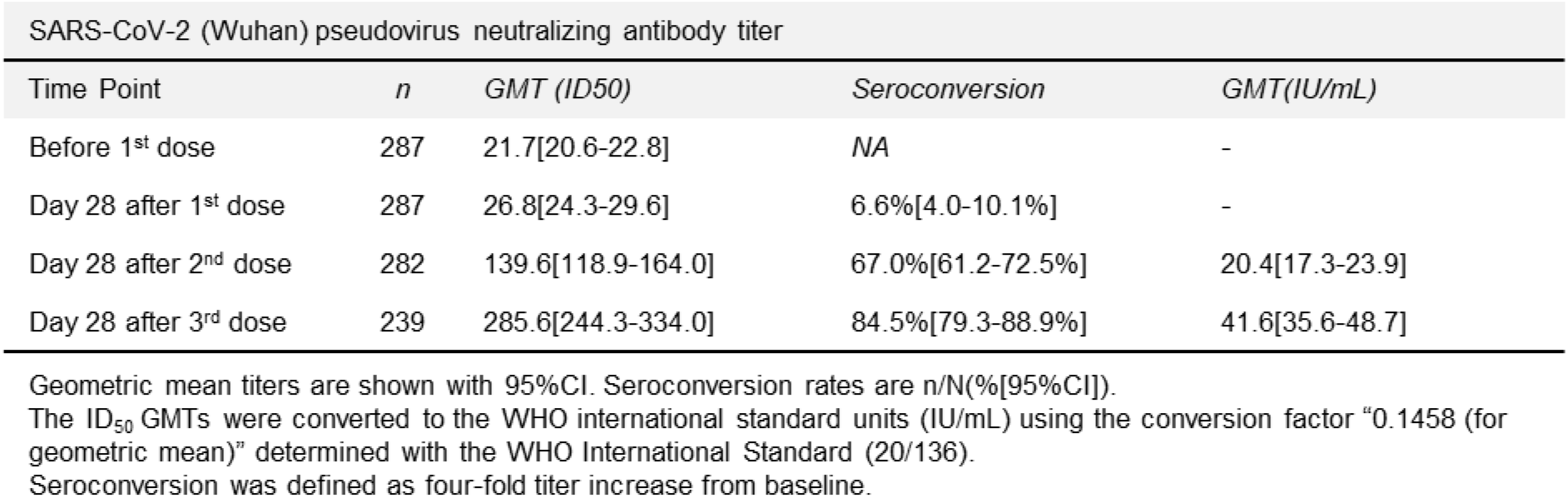
Geometric mean titers and seroconversion rate to SARS-CoV-2 (Wuhan) pseudovirus induced before first dose and after each dose of KD-414.

Analyses of neutralization GMTs by age subgroups of 18 - 40 years, 41 - 64 years, and &#x2265; 65 years showed that the values were 199.9 (163.6 - 244.4), 103.2 (79.9 - 133.2), and 50.7 (26.5 - 97.1), respectively, 28 days after the second vaccination, and 327.6 (269.8 – 397.9), 272.2 (211.5 - 350.4), and 128.0 (51.6 - 317.7), respectively, 28 days after the third vaccination. (Table 6)

**Table 6:** Geometric mean titers, seroconversion rate, and geometric mean titers converted to the WHO international standard units (IU/mL) induced after second and third doses of KD-414.

|  | Day 28 after 2<sup>nd</sup> dose |  |  |  | Day 28 after 3<sup>rd</sup> dose |  |  |  |
| --- | --- | --- | --- | --- | --- | --- | --- | --- |
|  | *n* | GMT (ID<sub>50</sub>) | Seroconversion rate | GMT (IU/mL) | *n* | GMT (ID<sub>50</sub>) | Seroconversion rate | GMT (IU/mL) |
| 18-40 years | 144 | 199.9[163.6-244.4] | 80.6%[73.1-86.7%] | 29.1[23.8-35.6] | 119 | 327.6[269.8-397.9] | 90.8%[84.1-95.3%] | 47.8[39.3-58.0] |
| 41-64 years | 124 | 103.2[79.9-133.2] | 54.8%[45.7-63.8%] | 15.0[11.6-19.4] | 106 | 272.2[211.5-350.4] | 80.2%[71.3-87.3%] | 39.7[30.8-51.1] |
| ≥65 years | 14 | 50.7[26.5-97.1] | 35.7%[12.8-64.9%] | 7.4[3.9-14.2] | 14 | 128.0[51.6-317.7] | 64.3%[35.1-87.2%] | 18.7[7.5-46.3] |

Geometric mean titers are shown with 95%CI. Seroconversion rates are n/N(%[95%CI]).
The ID<sub>50</sub> GMTs were converted to the WHO international standard units (IU/mL) using the conversion factor "0.1458 (for geometric mean)" determined with the WHO International Standard (20/136).
Seroconversion was defined as four-fold titer increase from baseline.
Geometric mean titers are shown with 95%CI. Seroconversion rates are n/N(%[95%CI]). The ID<sub>50</sub> GMTs were converted to the WHO international standard units (IU/mL) using the conversion factor "0.1458 (for geometric mean)" determined with the WHO International Standard (20/136). Seroconversion was defined as four-fold titer increase from baseline.

## Discussion

When intramuscularly administered three times to healthy Japanese participants aged 18 years or older who had no history of both COVID-19 and COVID-19 vaccination including unapproved vaccines, a favorable safety profile of KD-414 (10 μg/dose) was shown. The neutralizing antibody titers against pseudovirus SARS-CoV-2 (Wuhan) increased after the third vaccination. The most frequent adverse reaction was injection site pain, followed by relatively frequent adverse reactions such as malaise, headache, injection site erythema, myalgia, and injection site induration. Most adverse reactions were characterized as grade 1 or 2. A total of 11 grade 3 adverse reactions that prevented daily activities were reported in nine participants, and nine adverse reactions that led to the discontinuation of the following vaccination were reported in seven participants. Many grade 3 adverse reactions and adverse reactions leading to discontinuation of the following vaccination have resolved or have become less severe, and some are still under the follow-up observation. No deaths or serious adverse events with causal relationships were reported. The incidences of solicited local adverse reactions and fever after the third vaccination with KD-414 were similar to those of approved inactivated vaccines used worldwide, mainly in China (BBIBP-CorV, CoronaVac) ^15, 16^, and suggesting that the incidence may be relatively lower than those of approved mRNA vaccines which are currently mainly being used in Japan (Comirnaty, Spikevax) ^3, 4^, which were registered in the WHO Emergency Use Listing Procedure (EUL) ^1^.

In addition, no noteworthy trends were observed in the types and incidences of adverse reactions depending on the number of vaccinations of KD-414. In case that periodical COVID-19 vaccinations will be needed in the future, it should be more favorable for public health to be able to choose useful vaccines produced by several modalities having the profiles like low incidence of adverse reactions and no increase of adverse reactions depending on number of dosing. Since the number of enrolled participants with underlying diseases predetermined in this study protocol was small, further investigation on the safety profile of KD-414 in the specific population is necessary. Although the data is limited in this study, since no particular trend was observed in the incidence of adverse reactions depending on the presence or absence of underlying diseases, KD-414 is expected to be safe for that persons with underlying disease. Currently in Japan, only mRNA vaccine is available for children under five years of age. In addition, approved mRNA vaccines can be administered to pregnant women only when the benefits outweigh their risks. On the other hand, KD-414 has typically demonstrated relatively few adverse reactions, including those of grade 3 or higher. In this sense, KD-414 is expected to be more safely for children and pregnant women. In the future, it is necessary to evaluate safety of KD-414 in the aforementioned populations.

For immunogenicity, the product H (10 μg/dose) of KD-414 that induced the highest neutralizing antibody titers was selected for the subsequent phases based on the results after the second vaccination of KD-414 in the phase 1/2 study. However, although the preventive effect of the novel corona vaccine against the onset has been shown to correlate highly with the neutralizing antibody level, the threshold of the neutralizing antibody titers required for prevention has not yet been established. Therefore, it is important to evaluate the administration regimen of the three doses of KD-414, in which a higher neutralizing antibody titer can be expected. Therefore, the safety and immunogenicity of the three doses were evaluated in a phase 2/3 study using the administration method in which the third dose is administered three months after the second dose as the primary series. The results showed that GMTs increased by approximately two-folds 28 days after the third investigational product vaccination as compared with that 28 days after the second vaccination, and the extent of increase was similar to that observed after the third vaccination at 6 months after the second vaccination in the phase 1/2 study ^12^. The persistence of neutralizing antibody titers after the third vaccination is currently being evaluated in a phase 2/3 study. Analysis of GMTs by age group has shown that the tendency of reduction in the neutralizing antibody titers in the elderly was observed with KD-414, similar to the previously approved vaccines for SARS-CoV-2 ^17, 18^.

Currently, no surrogate marker has been established to evaluate the vaccine efficacy against COVID-19, and the method of measurement of neutralizing antibody titers differs among studies. Therefore, it is difficult to directly compare its efficacy with that of other vaccines. With reference to the literature that reported the vaccine efficacy (VE) of Vaxzevria (the active comparator in the currently ongoing pivotal phase 3 study examining KD-414) against the original strain, which was estimated from the neutralizing antibody titer level ^19^, we obtained the international standard units (IU) converted value of neutralization GMTs against pseudovirus SARS-CoV-2 (Wuhan) using the same method as in the literature, and attempted to estimate the VE of KD-414 against the original strain of SARS-CoV-2 after vaccination. The results shows that the KD-414 IU converted values of GMTs after the second and third vaccination in participants aged 18 - 40 years were 29.1 IU/mL and 47.8 IU/mL, respectively (Table 6). VE was estimated to be about 70% to 80%, respectively, suggesting that the efficacy of KD-414 could be comparable to that of the approved vaccines. In the future, based on the idea of rebalancing proposed by ICMRA ^20^, the efficacy of KD-414 is planned to be confirmed by verifying the superiority of immunogenicity to that of Vaxzevria in the ongoing pivotal phase 3 study.

In addition, it is necessary to examine the cross-reactivity of neutralizing antibody responses elicited by KD-414 vaccination against SARS-CoV-2 variants, and SARS-CoV-2 -specific cellular immune response. As a limitation of this study, the elevation of antibody titers as a results of asymptomatic infection was not fully excluded under the COVID-19 outbreak status. In addition, the cellular immune response by KD-414 vaccination will be evaluated in the currently ongoing phase 3 study in due course. The cross-reactivity of neutralizing antibody responses to SARS-CoV-2 variants and the incidence rate of COVID-19 infection after KD-414 dosing are currently being accumulated.

The aforementioned reports support the progress of the next phase of clinical studies on KD-414. The three-dose regimen with KD-414 as the primary vaccination is expected to provide good safety and induce high neutralizing antibody titers, especially in the age group of 18 to 40 years. Currently, phase 3 (jRCT2031210679) trial with adults aged 18 to 40 years is ongoing to verify the efficacy of three vaccinations with KD-414 as the primary vaccination. In addition, a phase 2/3 study (jRCT2031220032) in children aged &#x2265; 6 months to < 18 years is ongoing to evaluate the safety and immunogenicity of KD-414.

## Data Availability

All data produced in the present study are available upon reasonable request to the authors

## ACKNOWLEDGEMENT

This research was supported by Japan Agency for Medical Research and Development (AMED) under Grant Number JP21nf0101622 and Ministry of Health, Labour and Welfare.

We thank the Institute of Medical Science, the University of Tokyo which provided a virus strain (SARS-CoV-2/UT-HPCo-038/Human/2020/Tokyo).

We also thank all the participants who volunteered for this study.

## AUTHOR CONTRIBUTIONS

KK, KI, MT, and YM designed and coordinated this clinical trial. HN and YO advised technical expertise. KS and ME managed the investigational drug manufacturing and the neutralizing antibody assay. SN, KH, KA, YS, and YI managed and analyzed the data of this study. KU contributed to the clinical trial as a medical expert. KK wrote the first draft manuscript, and YS and TM wrote, reviewed and edited. All authors reviewed and approved the final version. All authors had full access to all the data in the study and had final responsibility for the decision to submit for publication.

